# The Adverse Event Atlas and Signal Consensus Index: A Multi-Source Pharmacovigilance Platform

**DOI:** 10.64898/2026.05.05.26352239

**Authors:** Andreas Bentsen

## Abstract

**Background:** Post-market pharmacovigilance relies predominantly on single-database disproportionality analysis of spontaneous adverse event reports, which lacks corroboration across independent evidence streams and cannot integrate randomised trial evidence. No publicly accessible platform has previously combined European national pharmacovigilance registries, the US FDA Adverse Event Reporting System (FAERS), and clinical trial meta-analyses into a unified, continuously scored signal detection framework.

**Methods:** We describe the Signal Consensus Index (SCI), a composite 0-100 pharmacovigilance signal score integrating disproportionality evidence from the Danish National Pharmacovigilance Database, the UK MHRA Yellow Card scheme, and FAERS, with DerSimonian-Laird meta-analytic risk ratios from ClinicalTrials.gov, across 6,905,874 drug-adverse event pairs. Each source contributes a continuous score derived from the lower bounds of three complementary disproportionality metrics (ROR, PRR, IC025) for spontaneous reporting sources, and from the pooled risk ratio lower confidence bound for clinical trials. The SCI is publicly accessible via the Adverse Event Atlas (aeatlas.com). We report reference set validation against the EU-ADR reference standard, a single-source comparison with discordance characterisation, temporal stability analysis across eight cumulative data windows (2015-2023), and a weight sensitivity analysis across seven pre-specified weighting schemes.

**Results:** The SCI generated 129,176 Moderate-or-Strong signals (SCI ≥ 50, confidence ≥ 50) and 7,290 Strong signals (SCI ≥ 70, confidence ≥ 70). Reference set validation against 88 classifiable drug-event pairs (44 positive controls, 44 negative controls) yielded 18 true positives, 0 false positives, 44 true negatives, and 26 false negatives (sensitivity 40.9%, specificity 100.0%, PPV 100.0%, NPV 62.9%). Zero false positives were observed across all 44 classifiable negative controls, with five false negatives attributable to the confidence gate correctly suppressing single-source signals pending multi-source corroboration. Single-source comparison demonstrated that FAERS alone generated 1,438,246 disproportionality signals, of which 94.8% were not confirmed by the SCI, while 54,184 SCI-detected signals were absent from FAERS, of which 8.3% involved drugs absent from the US reporting system. Discordance analysis showed that 99.8% of Danish non-confirmation reflected data availability constraints. Temporal stability was high: 98.5% of pairs received identical classifications across all seven weight scenarios, and 57.0% of final Strong signals were already detectable as Moderate or Strong in the earliest data window (2015-2016). Strong classifications were stable across weight scenarios (94.0% of Strong observations remaining Strong).

**Conclusions:** The SCI provides a transparent, openly accessible framework for cross-source pharmacovigilance signal prioritisation with 100% specificity and PPV against an established reference standard and stable classifications across weighting schemes. Progressive signal emergence through the Moderate tier supports its use as an early detection layer. The platform is available at aeatlas.com.

## Introduction

Post-market pharmacovigilance depends on the early and reliable detection of adverse drug reactions after medicines enter routine clinical use. The cornerstone of this process is spontaneous reporting, the voluntary submission of suspected adverse event reports by healthcare professionals and patients to national and international pharmacovigilance systems. Statistical disproportionality analysis of spontaneous reports has become the standard first-pass signal detection approach, enabling systematic surveillance across millions of drug-event pairs [1,2]. Methods including the Reporting Odds Ratio, Proportional Reporting Ratio, and Bayesian Information Component have been extensively studied and are applied routinely by regulators [9] including the European Medicines Agency, the US Food and Drug Administration, and national pharmacovigilance centres worldwide [3,4].

Despite the utility of disproportionality analysis, single-source signal detection has well-recognised limitations. Spontaneous reporting systems capture events that clinicians and patients choose to report, introducing reporting biases that vary by drug, indication, reporting culture, and regulatory jurisdiction. Database-specific characteristics, including size, therapeutic coverage, population demographics, and reporting incentive structures, substantially influence the performance of any signal detection algorithm [4]. The landmark comparison by Candore et al. demonstrated that no single algorithm consistently outperforms others across databases, and that achievable sensitivity and precision vary substantially with database characteristics rather than with the statistical method applied [4]. A signal detected in one database may reflect a local reporting artefact, an indication-specific confound, or a genuine pharmacological association, and a single source cannot reliably distinguish between these possibilities.

The logical response to this limitation is cross-source corroboration: requiring that a signal be detected independently across multiple data systems before it is prioritised for clinical review. Multi-database pharmacovigilance has been pursued through several frameworks, most notably the IMI PROTECT consortium, which evaluated signal detection performance across European regulatory and industry databases [5], and the EU-ADR project, which combined electronic healthcare records across six European countries for active surveillance [6]. These initiatives demonstrated that cross-database signal detection improves both sensitivity and specificity relative to any single source. However, existing multi-database approaches have predominantly compared signal detection outputs in parallel rather than combining them into a single composite score, and have generally relied on spontaneous reporting systems alone without integrating clinical trial evidence. Furthermore, no publicly available platform has implemented cross-source signal detection across both European national registries and the US FAERS system simultaneously, incorporating clinical trial data as a fourth independent evidence stream.

Clinical trial data offer a fundamentally different evidence type for pharmacovigilance. Randomised trials provide comparative risk estimates with denominators, enabling risk ratio quantification that spontaneous reporting cannot support. While individual trials are typically underpowered for rare adverse events, meta-analysis of trial-level arm data across registered studies can generate pooled estimates for systematic integration with spontaneous reporting signals [7]. The integration of clinical trial results from ClinicalTrials.gov into pharmacovigilance signal detection has not been operationalised at scale in prior published frameworks.

We describe the Signal Consensus Index (SCI), a composite 0-100 pharmacovigilance signal score that integrates disproportionality evidence from two European national spontaneous reporting systems, the Danish National Pharmacovigilance Database and the UK MHRA Yellow Card scheme, with FAERS and with meta-analysed clinical trial data from ClinicalTrials.gov. The SCI is implemented within the Adverse Event Atlas (aeatlas.com), a publicly accessible cross-source pharmacovigilance platform covering 6,905,874 drug-adverse event pairs. We describe the SCI construction, validation framework, and performance characteristics, and present a reference set validation against the EU-ADR reference standard (Coloma et al. 2013), a single-source comparison with discordance characterisation, and a weight sensitivity analysis demonstrating robustness of signal classifications across plausible weighting schemes.

## Methods

### 1.1 Study design and platform overview

The Adverse Event Atlas (aeatlas.com) is a cross-source pharmacovigilance platform that integrates four independent evidence streams to identify and prioritise drug safety signals. The platform applies disproportionality analysis and meta-analytic methods to each source and combines source-specific signal scores into a composite Signal Consensus Index (SCI). The SCI is designed to detect signals that are concordant across independent regulatory jurisdictions and data types, thereby reducing false positive signals arising from reporting artefacts, indication confounding, or database-specific biases inherent in any single spontaneous reporting system.

### 1.2 Data sources

Four primary data sources contributed independent evidence streams, each operating under distinct regulatory frameworks, reporting cultures, and patient populations.

#### 1.2.1 Danish National Pharmacovigilance Database (DK)

The Danish Medicines Agency operates a population-based pharmacovigilance registry with near-universal prescription coverage through linkage to national prescription records. Adverse drug reaction reports are submitted by healthcare professionals and patients and coded using the Medical Dictionary for Regulatory Activities (MedDRA) [10] terminology. Data covering 2015 through 2023 were included.

#### 1.2.2 UK MHRA Yellow Card Scheme (UK)

The Medicines and Healthcare products Regulatory Agency (MHRA) Yellow Card scheme is the United Kingdom’s national spontaneous adverse drug reaction reporting system, established in 1964. Reports are submitted by healthcare professionals, patients, and carers and coded using MedDRA terminology. Following the United Kingdom’s exit from the European Union, the Yellow Card scheme operates as an independent national system providing a distinct European regulatory perspective. Data covering Q1 2015 through Q4 2025 were included.

#### 1.2.3 FDA Adverse Event Reporting System (FAERS)

The FDA Adverse Event Reporting System is the United States Food and Drug Administration’s post-market spontaneous reporting database, capturing adverse event reports from clinicians, patients, and manufacturers submitted from the US and internationally. Quarterly data files from Q1 2016 through Q4 2025 were included. For each case identifier, only the most recent version was retained to avoid duplicate counting.

#### 1.2.4 ClinicalTrials.gov via AACT database (CT)

Structured results from registered clinical trials were accessed via the Aggregate Analysis of ClinicalTrials.gov (AACT) database. Only trials with posted results were included. Arm-level adverse event counts and participants at risk were extracted for drug-containing interventions. Treatment and control arms were classified using predefined heuristics based on arm title normalisation. Clinical trial data provides comparative risk estimates with denominators, complementing spontaneous reporting sources which lack denominator information.

### 1.3 Disproportionality analyses

For spontaneous reporting sources, disproportionality was assessed using a standard 2×2 contingency table framework comparing reports of a given outcome for a specific drug against all other drugs in each database. The following metrics were calculated: Reporting Odds Ratio (ROR) with 95% confidence interval (CI); Proportional Reporting Ratio (PRR) with 95% CI; and Bayesian Information Component (IC) with lower 95% bound (IC025). All three spontaneous reporting sources use the same disproportionality framework, with all three metrics (ROR, PRR, IC025) contributing to a weighted composite source score as described in Section 1.5.1. For clinical trials, the DerSimonian-Laird pooled risk ratio lower CI bound is used directly. The SCI does not apply a binary signal threshold within individual sources; source scores are continuous. For descriptive reporting and the single-source comparison in the validation analyses, a source was classified as signal-positive when the source score exceeded 50, equivalent to a lower CI bound exceeding the null with meaningful effect size.

### 1.4 Clinical trial meta-analysis

Per-study risk ratios (RR) and risk differences were calculated from arm-level counts. Studies were pooled using a DerSimonian-Laird random-effects model with I^2^ heterogeneity assessment. A continuity correction of 0.5 was applied to zero-event cells; however, when all events occurred in the treatment arm across contributing studies, the meta-analysis was classified as unscorable and the clinical trial source was excluded from the composite for that pair. A minimum of two qualifying trials with at least 50 participants at risk was required for the clinical trial component to contribute to the SCI.

### 1.5 Signal Consensus Index

#### 1.5.1 Composite score construction

The Signal Consensus Index (SCI) is a composite 0-100 score combining source-specific signal components using equal weights across contributing sources. The SCI is computed as the arithmetic mean of available source scores: SCI = mean(source scores). Where a source did not meet minimum data thresholds it was excluded, and the composite was computed as the mean of the remaining contributing sources. All three spontaneous reporting sources (DK, UK, FAERS) use the same disproportionality framework and scoring formula, reflecting that all three are spontaneous reporting systems analysed without exposure denominators. Three complementary disproportionality metrics are combined using fixed weights: ROR lower 95% CI bound (weight 0.40), PRR lower 95% CI bound (weight 0.35), and IC025 (weight 0.25). ROR is weighted most heavily for its interpretability; PRR captures a related but non-identical disproportionality dimension; and IC025 provides Bayesian stabilisation for sparse cells. Clinical trial data, representing randomised comparative evidence, contribute a meta-analytic risk ratio as the most causally interpretable evidence type available. The SCI therefore applies a consistent estimand-aware design: spontaneous reporting sources share a common disproportionality estimand appropriate for systems without reliable denominators, while clinical trials contribute a randomised comparative estimand. This distinction is intentional: each estimator is chosen for alignment with the structure of its data source. Across evidence streams, the resulting source scores are not measuring the same underlying quantity. Rather, each score reflects the strength of evidence from that source that a pharmacological association exists between the drug and adverse event. The SCI aggregates these independent lines of evidence into a composite prioritisation score, on the premise that concordance across methodologically independent sources provides stronger grounds for signal prioritisation than any single source alone, regardless of whether the sources are measuring directly comparable quantities. This framework is analogous to regulatory signal assessment practice, where spontaneous reporting signals, clinical trial data, and epidemiological evidence are routinely triangulated without requiring formal commensurability. The SCI is designed as a composite prioritisation score for ranking drug-adverse event pairs by cross-source evidence concordance, not as a unified causal effect estimate. Equal weighting across sources at the aggregation stage reflects the absence of a prior basis for privileging one regulatory jurisdiction over another; the weight sensitivity analysis (Section 1.6.4) demonstrates that SCI classifications are robust to this choice. A confidence score (0-100) reflects source coverage and is computed as: confidence = 100 × (sources used / 4), where sources used is the count of sources contributing a valid score to the composite. A confidence of 100 indicates all four sources contributed; 75 indicates three sources; 50 indicates two sources.

Each source score is derived from the lower confidence bound of the primary effect estimate, scaled logarithmically to a 0-100 range: source_score = 100 × clamp(ln(effect_LCL) / ln(3), 0, 1). A lower confidence bound of 3 (tripling of risk relative to background) corresponds to a full source score of 100; a lower confidence bound at or below 1 (null or protective direction) yields a source score of 0, with proportional log-linear scaling for intermediate effect sizes. The choice of ln(3) as the scaling reference is a heuristic normalisation: a lower confidence bound of 3 represents a strong and clinically meaningful signal in spontaneous reporting practice, and using it as the ceiling anchors the scale to a pharmacovigilance-relevant threshold rather than an arbitrary statistical cutoff. The logarithmic transformation reflects the diminishing discriminative value of effect size at very high magnitudes. This scaling is intended to produce comparable rank orderings across sources, not to equate the effect estimates themselves.

#### 1.5.2 Source inclusion gating

No minimum data threshold is applied to DK, UK, or FAERS; these sources contribute a score whenever a computable disproportionality estimate is available, with a source score of 0 when the lower confidence bound is at or below the null. The clinical trial source required a minimum of two qualifying trials and at least 50 participants at risk. There is no binary inclusion or exclusion for any spontaneous reporting source beyond the requirement of a computable estimate.

#### 1.5.3 Signal classification

SCI results were classified into three tiers. Strong: SCI ≥ 70 and confidence ≥ 70. Moderate: SCI ≥ 50 and confidence ≥ 50, but not meeting Strong criteria. Weak/No signal: SCI < 50 or confidence < 50. Classification is hierarchical: Strong is evaluated first, then Moderate among remaining pairs, with all others classified as Weak. The Strong classification requires both high signal magnitude and sufficient cross-source confidence (at least three of four sources contributing, yielding confidence ≥ 75), ensuring that signals from only one or two sources are not classified as Strong regardless of effect size.

### 1.6 Validation

#### 1.6.1 Reference set validation

Reference set validation was conducted using the EU-ADR reference standard described by Coloma et al. (2013), which comprises 44 positive drug-adverse event associations and 50 negative controls across ten clinically important adverse event types: acute liver injury, acute myocardial infarction, acute renal failure, anaphylactic shock, bullous eruptions, cardiac valve fibrosis, neutropenia/agranulocytosis, aplastic anaemia/pancytopenia, rhabdomyolysis, and upper gastrointestinal bleeding. Positive associations were established through systematic MEDLINE review, drug product label verification, and independent expert adjudication (inter-rater kappa 0.83). Negative controls were verified to have no MEDLINE citations for the drug-event association, no mention in the drug product label, and no signal flagged in VigiBase [12] using Bayesian disproportionality analysis. This reference set was specifically designed for evaluation of spontaneous reporting and electronic healthcare record-based signal detection methods, making it directly applicable to the SCI validation context. Each of the 94 drug-event pairs was looked up against the SCI materialized view using exact drug name and MedDRA Preferred Term matching. Drug name variants were investigated where pairs were not found on the primary INN search, given the known cross-jurisdictional naming inconsistencies documented in Section 1.5. The SCI was classified as signal-positive at the Moderate-or-Strong threshold (SCI ≥ 50 and confidence ≥ 50) and sensitivity, specificity, positive predictive value (PPV), and negative predictive value (NPV) were calculated for the subset of pairs found in the Atlas.

#### 1.6.2 Single-source comparison and discordance analysis

To evaluate the added value of cross-source aggregation relative to single-source disproportionality analysis, the SCI was compared with the signal classification from each individual source (DK, UK, FAERS, CT) across all 6,905,874 drug-adverse event pairs in the Atlas database. For each pair, binary signal status was determined for the SCI (Moderate-or-Strong, defined as SCI ≥ 50 and confidence ≥ 50) and for each source (source score ≥ 50). This threshold captures all pairs where the SCI detects any signal, encompassing both the Moderate and Strong classification tiers. Agreement, concordant signal pairs, SCI-only signals, and source-only signals were enumerated. McNemar’s test assessed the significance of directional disagreement between the SCI and each source (all p < 2.2 × 10^−16^).

Source-specific discordance analysis was performed for all SCI-positive signals not confirmed by each individual source. Reasons for non-confirmation were categorised as: drug present with no report of the specific pair, pair sparse (below the hard inclusion gate for CT, or no computable estimate for PV sources), drug absent from the source entirely, score below the signal threshold (disproportionality estimate computable but source score < 50), and no estimate computable. For DK, a supplementary component analysis characterised discordant pairs by the number of positive disproportionality components (0, 1, 2, or all 3 of ROR/PRR/IC lower bounds exceeding the null), to assess whether discordance reflected true absence of signal or subthreshold signal in the same direction.

#### 1.6.3 Temporal stability analysis

To assess whether SCI classifications are stable as data accumulate over time, a temporal stability analysis was conducted across eight cumulative data windows spanning 2015 to 2023, defined at one-to two-year intervals: 2015-2016, 2015-2017, 2015-2018, 2015-2019, 2015-2020, 2015-2021, 2015-2022, and 2015-2023. For each window, source-specific disproportionality estimates and clinical trial meta-analyses were recomputed using only reports and trial data falling within the window period. SCI scores and classifications (Strong, Moderate, Weak) were then derived using the primary analysis weights and classification thresholds. Classification stability was assessed at pair level by comparing the classification assigned in each window against the final (2015-2023) classification. Signal emergence was characterised for the 7,330 pairs classified as Strong in the final window by tracking the proportion classified as Strong, Moderate, or Weak in each preceding window.

#### 1.6.4 Weight sensitivity analysis

To assess the robustness of SCI classifications to the choice of source weights, a pre-specified sensitivity analysis was conducted in which source weights were varied across seven alternative scenarios covering the principal plausible weighting philosophies (Table 5). Scenarios included equal weighting (25% each source), spontaneous reporting emphasis (UK and FAERS each 30%), clinical trial emphasis (CT 40%), European source emphasis (DK and UK each 30%), and individual source dominance for FAERS (45%), DK (40%), and UK (40%). The range of weights tested spanned 15-45% for any individual source, representing a wide variation around the primary analysis equal weighting (25% each source). For each scenario, SCI scores were recomputed using the same source exclusion and normalisation logic as the primary analysis, and the resulting classification (Strong, Moderate, Weak) was compared with the baseline. Classification stability was defined as receiving the same classification across all seven weight scenarios (perfectly stable) or in at least five of seven scenarios (mostly stable).

### 1.7 Ethical considerations and data availability

All data sources are publicly available or accessible through regulatory data sharing programmes. No individual patient data were accessed; all analyses were performed on aggregated summary statistics. The Adverse Event Atlas platform is publicly accessible at aeatlas.com and the SCI methodology is fully documented at aeatlas.com/wiki.

## Results

### 2.1 Reference set validation (Coloma EU-ADR)

Of 94 drug-event pairs from the EU-ADR reference set, 88 were found in the Adverse Event Atlas with a scorable SCI. The 6 pairs not found were all negative controls (cardiac fibrosis pairs for estradiol, fluvoxamine, furosemide, irbesartan, and methotrexate; and fluvastatin-aplastic anaemia); these were excluded from the confusion matrix per standard reference set methodology. The 88 classifiable pairs comprised 44 positive controls and 44 negative controls. Drug name fragmentation required synonym resolution for several pairs: amoxicillin clavulanate appears as amoxycillin in UK/DK databases and amoxicillin in FAERS (queried under both spellings), thiamazole as methimazole in FAERS, and glyceryl trinitrate as nitroglycerin in FAERS. Among the 88 classifiable pairs, the SCI yielded 18 true positives, 0 false positives, 44 true negatives, and 26 false negatives at the Moderate-or-Strong threshold (SCI ≥ 50 and confidence ≥ 50), corresponding to sensitivity 40.9%, specificity 100.0%, PPV 100.0%, and NPV 62.9% (Table 1). At the Strong threshold (SCI ≥ 70 and confidence ≥ 70), sensitivity was 18.2%, specificity 100.0%, and PPV 100.0%. Five of the 26 false negatives had SCI scores ≥ 50 but confidence scores of 25 (single contributing source), indicating that the cross-source confidence requirement correctly suppressed single-source signals where multi-source corroboration was absent (valdecoxib-acute myocardial infarction, ticlopidine-aplastic anaemia, sulfamethoxazole-Stevens-Johnson syndrome, thiamazole-agranulocytosis, heparin-gastrointestinal haemorrhage).

**Table 1.**
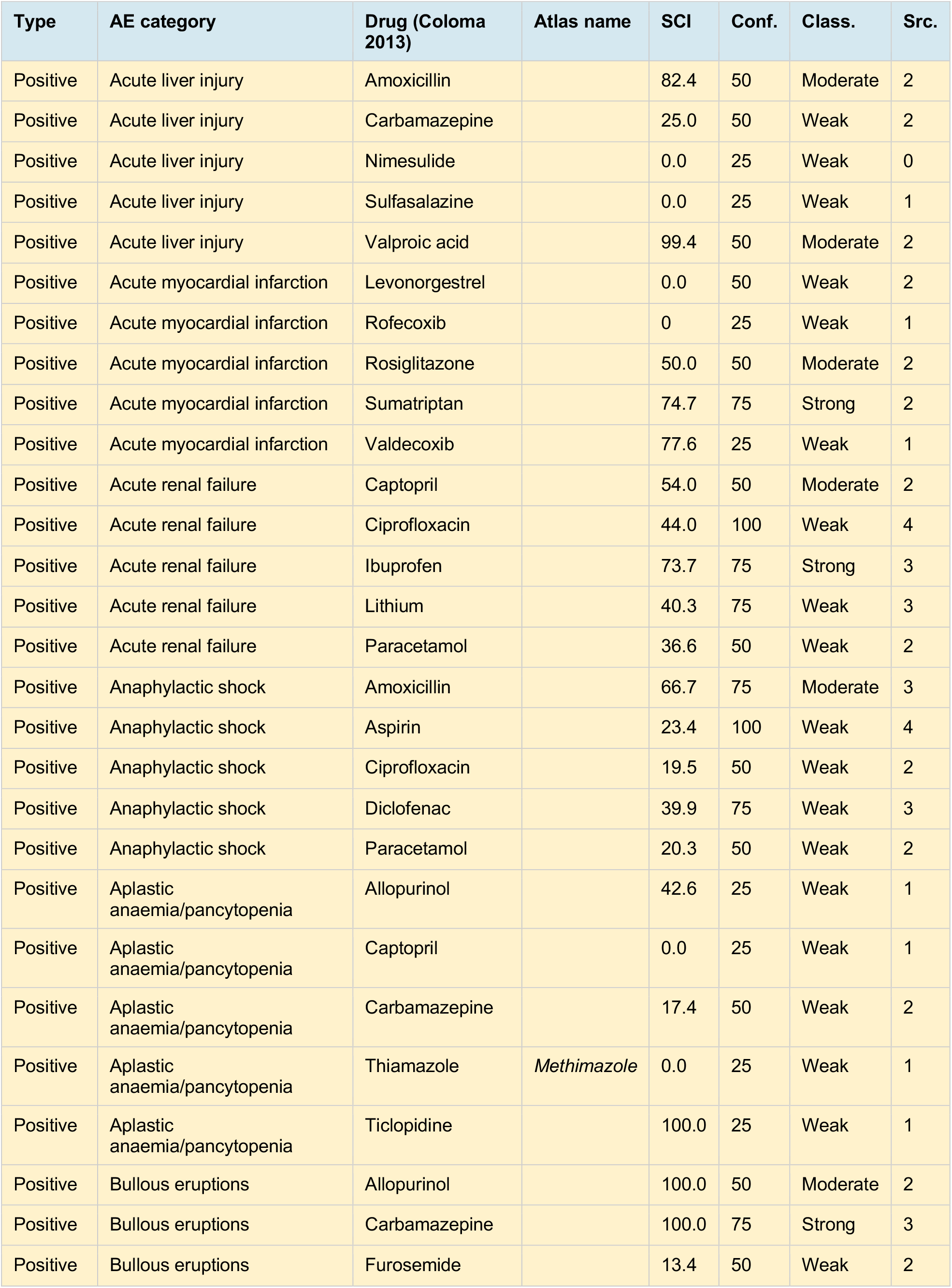
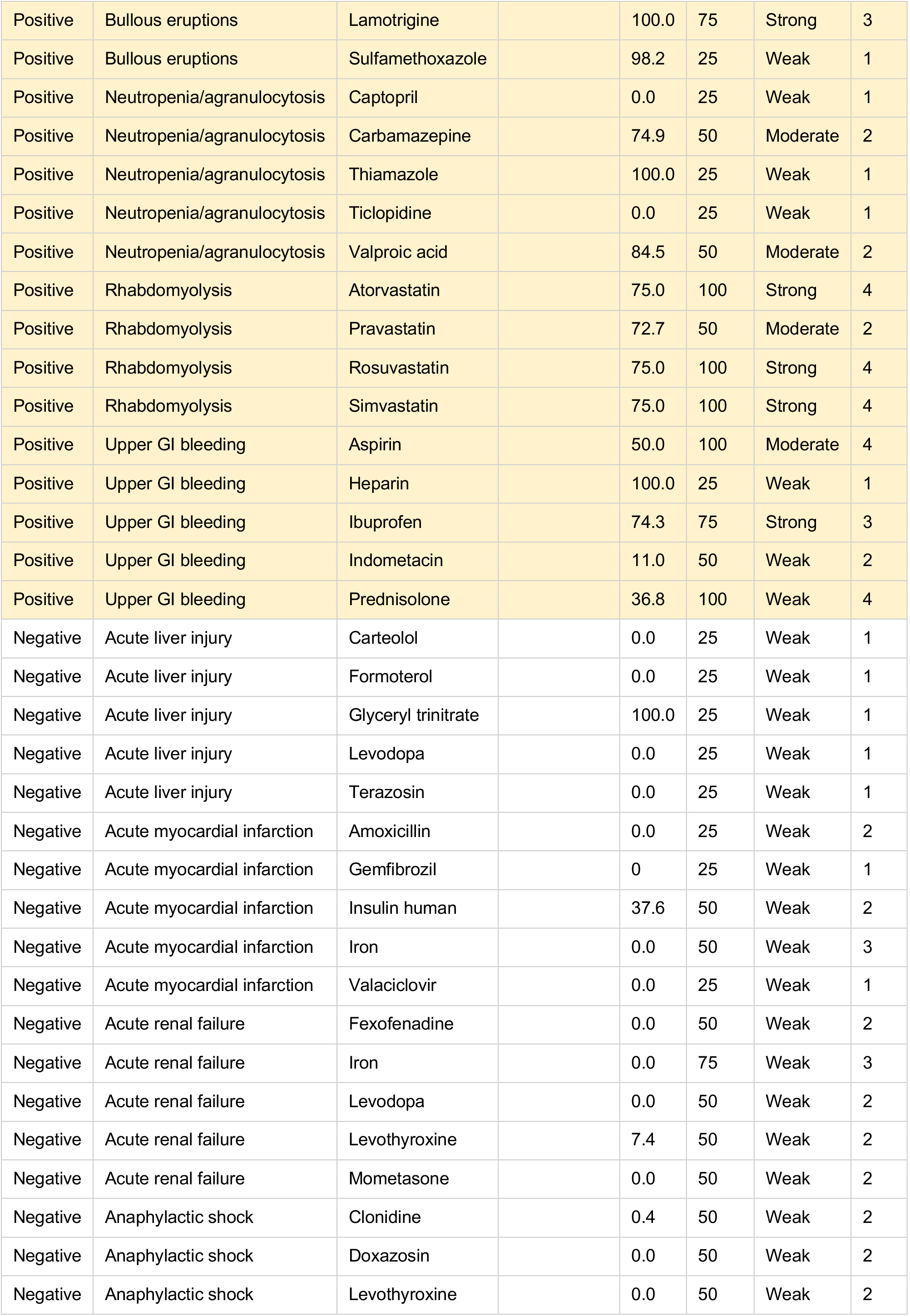
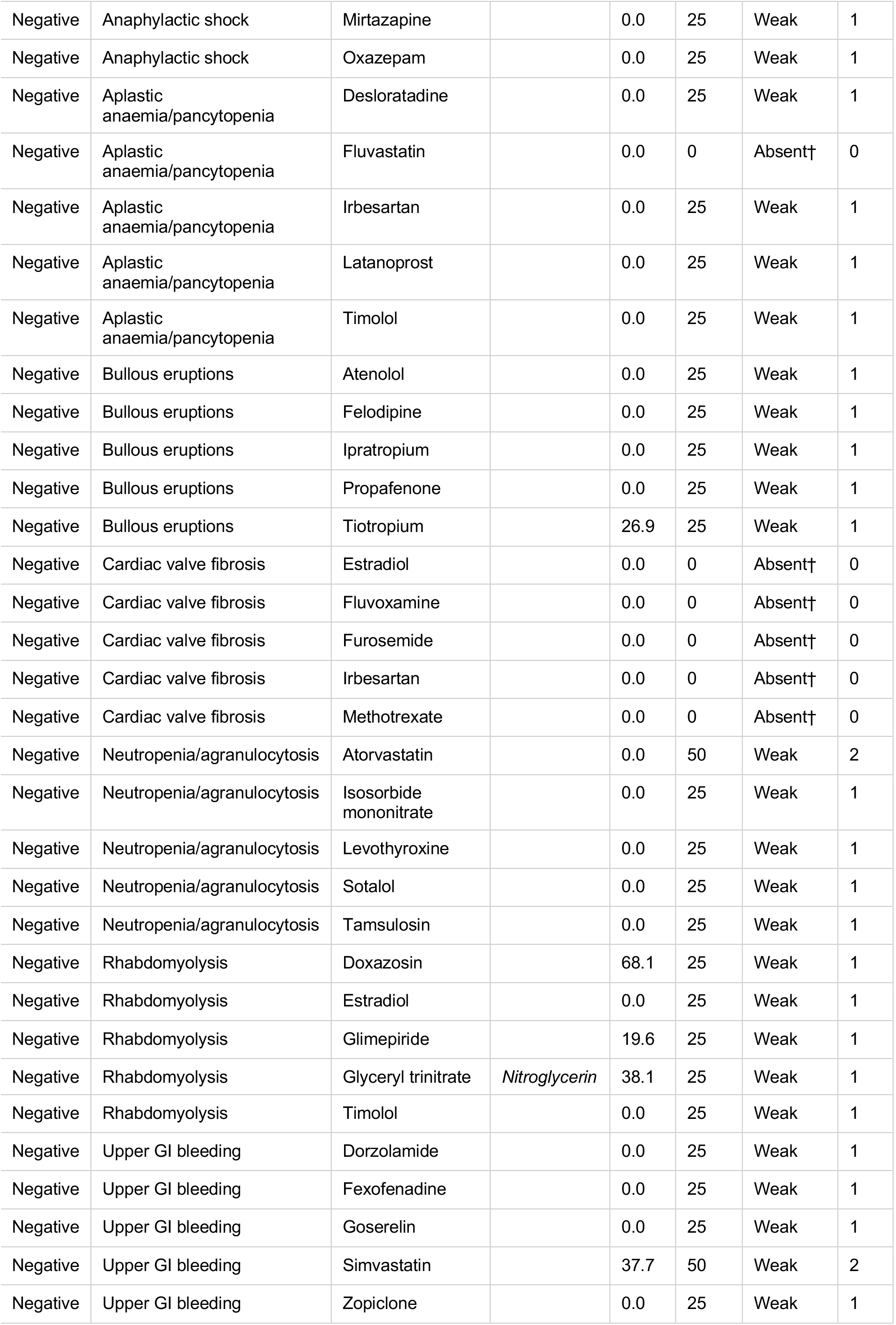
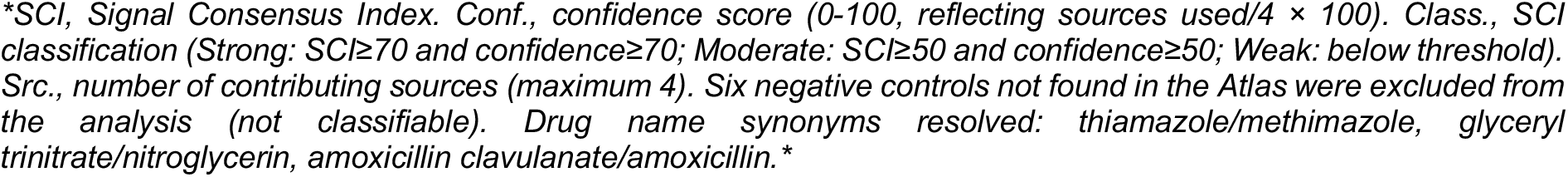
EU-ADR reference set validation (Coloma 2013): SCI scores for 88 classifiable drug-adverse event pairs (44 positive controls, 44 negative controls). Six negative controls not found in the Atlas were excluded from analysis (not classifiable).

The zero false positive rate across all 44 classifiable negative controls is a structural property of the cross-source confirmation requirement: no negative control pair achieved sufficient concordance across independent evidence streams to be classified as a signal, even where individual source scores were elevated. Five false negatives had SCI scores ≥ 50 but confidence scores of 25, reflecting single-source signals where multi-source corroboration was absent; the confidence gate correctly suppressed these to Weak rather than generating false Moderate classifications. The practical implication is that Moderate-classified signals warrant pharmacological attention when data availability constraints explain the confidence penalty, as the signal score itself exceeds the detection threshold.

Sensitivity was limited by two primary mechanisms. First, the confidence threshold requiring at least two contributing sources was the dominant driver: five false negatives had SCI scores ≥ 50 but confidence scores of 25, correctly suppressed to Weak pending multi-source corroboration (valdecoxib-acute myocardial infarction, ticlopidine-aplastic anaemia, sulfamethoxazole-Stevens-Johnson syndrome, thiamazole-agranulocytosis, heparin-gastrointestinal haemorrhage). Several further false negatives involved withdrawn drugs (rofecoxib, valdecoxib) or drugs with limited current reporting. Second, several established associations showed genuinely weak disproportionality signals across all available sources, reflecting inherent limitations of spontaneous reporting for high-background drug-AE pairs: aspirin-anaphylactic reaction (SCI 23.4) and prednisolone-gastrointestinal haemorrhage (SCI 36.8) both showed low effect estimates consistent with indication confounding and ubiquitous drug use suppressing disproportionality.

The 6 negative controls not found in the Atlas (five cardiac fibrosis pairs and fluvastatin-aplastic anaemia) were excluded from the confusion matrix per standard reference set methodology; pairs absent from the database cannot be classified and are therefore not interpretable as true negatives or false positives. SCI scores for all 88 classifiable pairs are presented in Table 1. Clinical trial data contributed no scorable estimates to any reference set pair, reflecting the predominance of older drug classes in the Coloma reference set and the rarity of the targeted adverse events in randomised trial populations.

### 2.2 Single-source comparison and discordance analysis

Across 6,905,874 drug-adverse event pairs evaluated, the SCI generated 129,176 Moderate-or-Strong signals (SCI ≥ 50 and confidence ≥ 50), of which 7,290 were classified as Strong (SCI ≥ 70 and confidence ≥ 70) and 121,886 as Moderate. For comparison, applying an equivalent score threshold of ≥ 50 to individual sources yielded 44,601 signals for the Danish source alone, 165,036 for the UK source alone, 1,438,246 for FAERS alone, and 1,268 for clinical trials alone (Figure 1, Table 2). Overall agreement between the SCI and each source was ≥ 98% for DK, UK, and CT, driven by the large majority of pairs being non-signals in every system; agreement with FAERS was substantially lower at 79.5%, reflecting the much larger volume of FAERS-only disproportionality signals that the SCI does not confirm. McNemar’s test confirmed statistically significant asymmetry in SCI-source disagreement for all four sources (all p < 2.2 × 10^−16^). The direction of asymmetry differs by source: for DK and CT the SCI detects substantially more signals than the source alone (SCI-only pairs predominate), for UK the source detects more pairs than the SCI (source-only pairs predominate), and for FAERS the source-only excess is extreme (1,363,254 FAERS-only pairs versus 54,184 SCI-only pairs). The SCI is therefore not merely a reflection of single-source classifications but an aggregation that sits between the most permissive and the most restrictive source-level thresholds.

**Figure 1.**
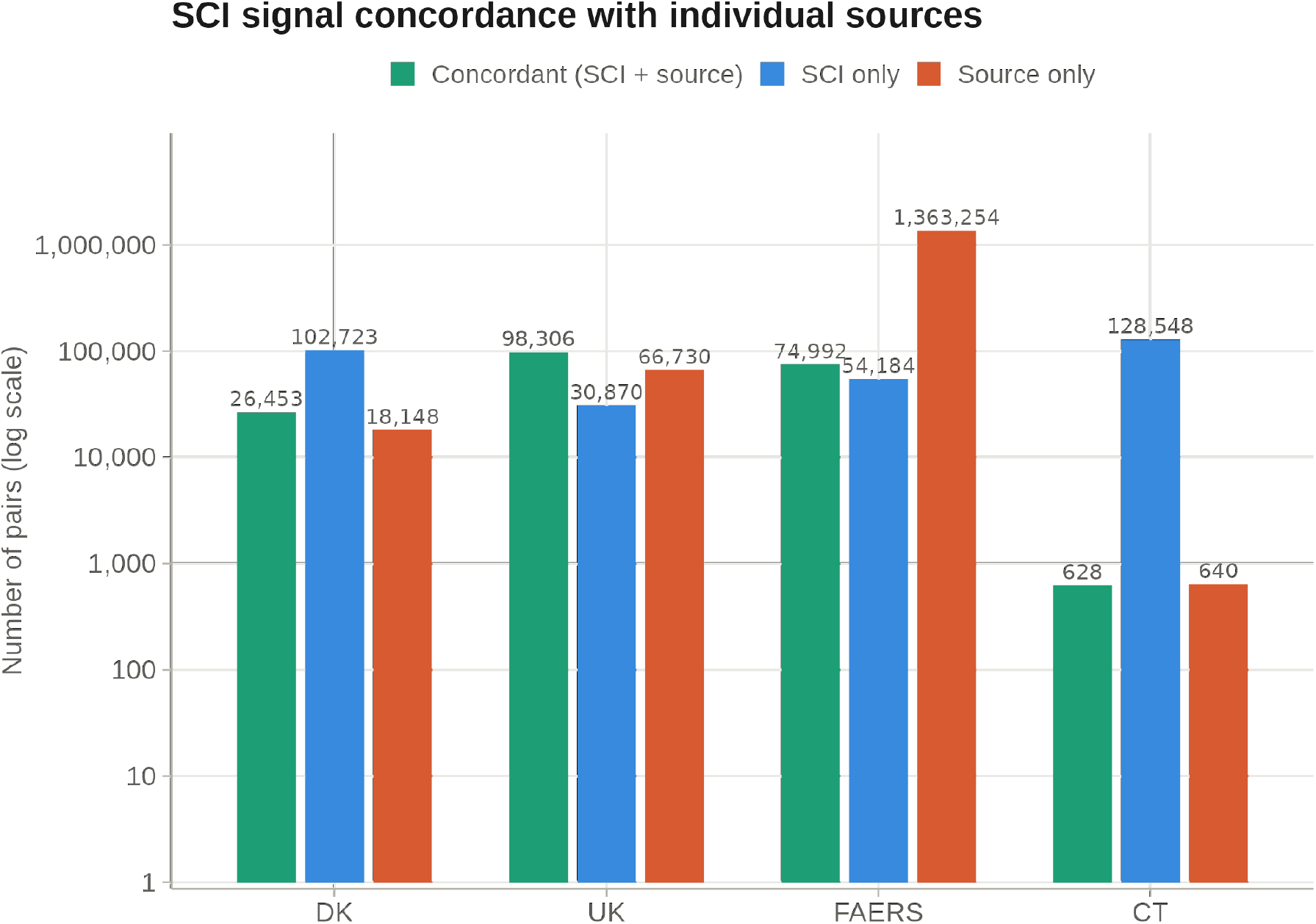
SCI signal concordance with individual sources across 6,905,874 drug-adverse event pairs (logarithmic scale). Bars show concordant signals (detected by both SCI and source), SCI-only signals (not confirmed by source), and source-only signals (not captured by SCI).

**Table 2.** Single-source comparison: SCI signal concordance with each individual data source across 6,905,874 drug-adverse event pairs.

|  | DK | UK | FAERS | CT |
| --- | --- | --- | --- | --- |
| Total SCI signals | 129,176 | 129,176 | 129,176 | 129,176 |
| Source signals (total) | 44,601 | 165,036 | 1,438,246 | 1,268 |
| Concordant (SCI + source) | 26,453 | 98,306 | 74,992 | 628 |
| SCI-only (not confirmed by source) | 102,723 | 30,870 | 54,184 | 128,548 |
| Source-only (not captured by SCI) | 18,148 | 66,730 | 1,363,254 | 640 |
| Overall agreement | 98.2% | 98.6% | 79.5% | 98.1% |
| SCI sensitivity vs. source | 59.3% | 59.6% | 5.2% | 49.5% |
| SCI specificity vs. source | 98.5% | 99.5% | 99.0% | 98.1% |
| SCI PPV vs. source | 20.5% | 76.1% | 58.1% | 0.5% |

DK, Danish National Pharmacovigilance Database; UK, MHRA Yellow Card; FAERS, FDA Adverse Event Reporting System; CT, ClinicalTrials.gov. SCI sensitivity: proportion of source-positive signals confirmed by the SCI. SCI specificity: proportion of source-negative pairs not flagged by the SCI. PPV: positive predictive value of SCI classification relative to each source.

The comparison with FAERS provided the most direct demonstration of cross-source filtering of spontaneous reporting volume. FAERS alone generated 1,438,246 signals at the ≥ 50 score threshold, of which 1,363,254 (94.8%) were not confirmed by the SCI at the Moderate-or-Strong tier, representing a substantial reduction of single-source disproportionality output to a cross-source corroborated subset. The 74,992 FAERS signals that were SCI-confirmed correspond to a positive predictive value of 58.1% of SCI Moderate-or-Strong signals being FAERS-positive, and conversely 5.2% of FAERS single-source signals being SCI-confirmed. Conversely, 54,184 SCI-detected signals were not detected by FAERS. Of these, 8.3% involved drugs absent from FAERS entirely, representing European-authorised medicines with limited US prescribing that a FAERS-only system would not detect; a further 11.3% involved drug-AE pairs with no reports in FAERS despite the drug being present; 24.5% had fewer than three FAERS cases; and 55.9% had FAERS disproportionality scores below the ≥ 50 threshold, reflecting weaker US reporting for associations driven primarily by European spontaneous reporting data.

Source-specific discordance analysis characterised why SCI-detected signals (Moderate-or-Strong) were not confirmed by each individual source (Figure 2, Table 3). For the 102,723 SCI-detected signals not confirmed by the Danish source, 99.8% were attributable to data availability constraints: 87.5% had no Danish reports for the specific drug-AE combination despite the drug being present in Danish data, 11.0% involved drugs absent from Danish data entirely, and 1.3% had fewer than three Danish cases. A further 0.02% had no computable disproportionality estimate. Only 205 pairs (0.2%) had a source score below the ≥ 50 threshold; among these, the median DK score was 25.9, the median ROR lower CI bound was 1.33, and all three disproportionality components were positive (ROR, PRR, and IC025 all exceeding the null), while FAERS (median score 94) and UK (median score 91) showed strong signals for the same pairs. These 205 pairs therefore represent SCI signals where DK data provides a subthreshold signal in the same direction as FAERS and UK — insufficient to independently confirm the SCI classification but not contradicting it. For the 30,870 SCI-positive signals not confirmed by the UK source, data availability dominated (93.7%): 70.5% had the drug present but no UK reports for the pair, 20.5% had fewer than three UK cases, and 2.7% involved drugs absent from UK data entirely; the remaining 6.3% had UK scores below the ≥ 50 threshold. For FAERS, 44.1% of the 54,184 discordant pairs reflected data availability constraints (24.5% pair sparse, 11.3% drug present but pair absent, 8.3% drug absent entirely), while 55.9% had FAERS scores below the threshold. For clinical trials, 94.6% of the 128,548 discordant pairs reflected data availability (66.9% drug present but no trial evidence for the specific pair, 23.0% drug absent entirely, 4.7% fewer than two qualifying trials or below the minimum at-risk threshold), and 5.4% had trial scores below the threshold. Characterisation of the discordant CT pairs confirmed that the dominant pattern is genuine absence of trial evidence rather than statistical underpowering: the median CT-discordant pair had zero qualifying studies, zero participants at risk, and zero trial arm-level events, with a median CT source score of 0. Under the updated SCI, no CT discordant pairs were classified as meta-analysis unscorable (trial evidence present but risk ratio uncomputable due to zero-event control arms); such pairs are now absorbed into the pair-sparse or subthreshold categories through the inclusion gate and score-of-zero logic. Across all four sources, discordance between the SCI and the individual source was therefore dominated by data availability and threshold effects rather than by conflicting pharmacological evidence.

**Figure 2.**
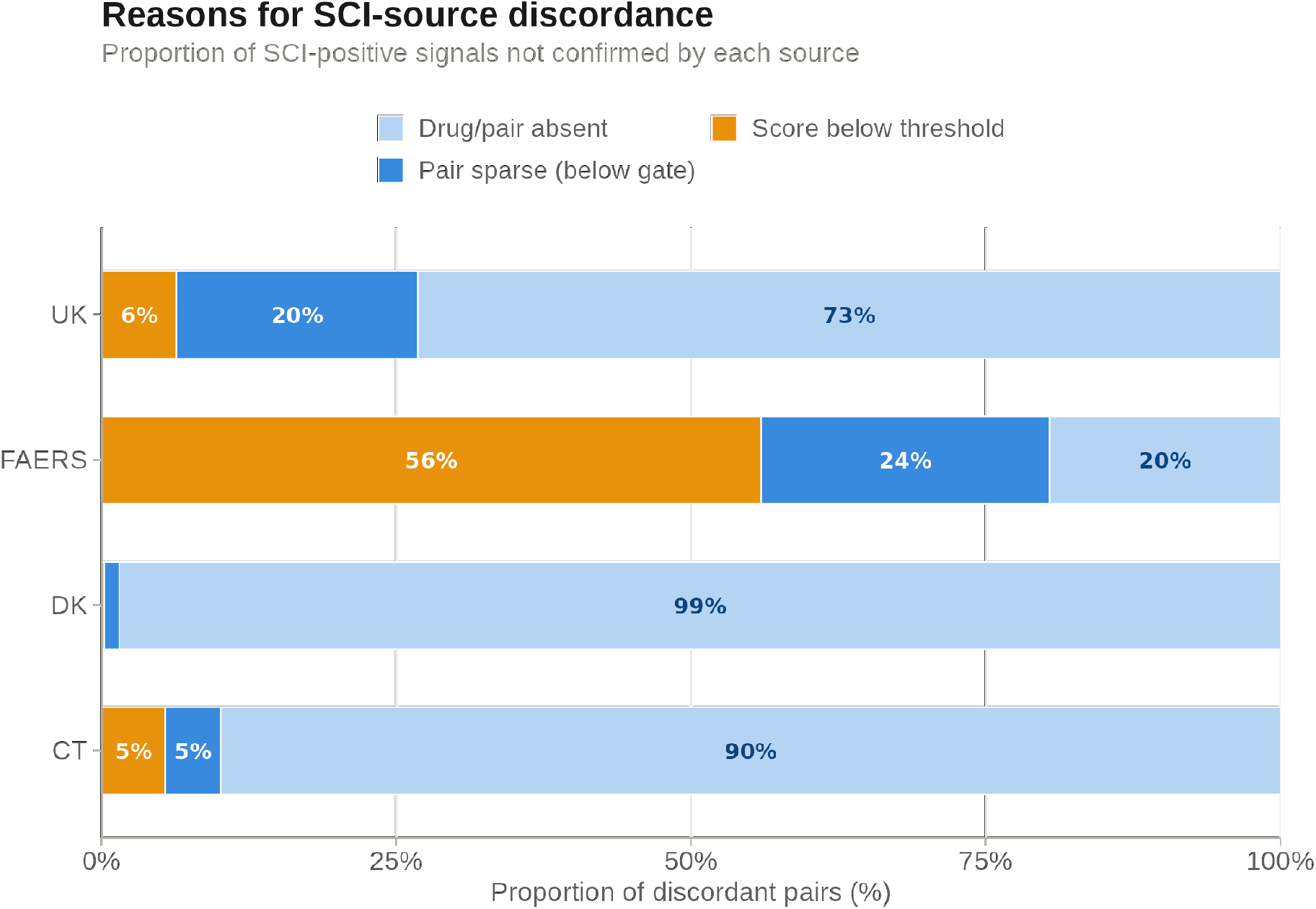
Reasons for non-confirmation of SCI-positive signals by each individual source, as a proportion of all discordant pairs. Discordance for DK, UK, and CT is dominated by data availability constraints (drug or pair absent from source, or below hard inclusion gate for CT); FAERS discordance is split between data availability (44%) and score threshold effects (56%). All three spontaneous reporting sources use the same disproportionality framework in SCI v6; no Mantel-Haenszel or case-count attenuation categories apply.

**Table 3.**
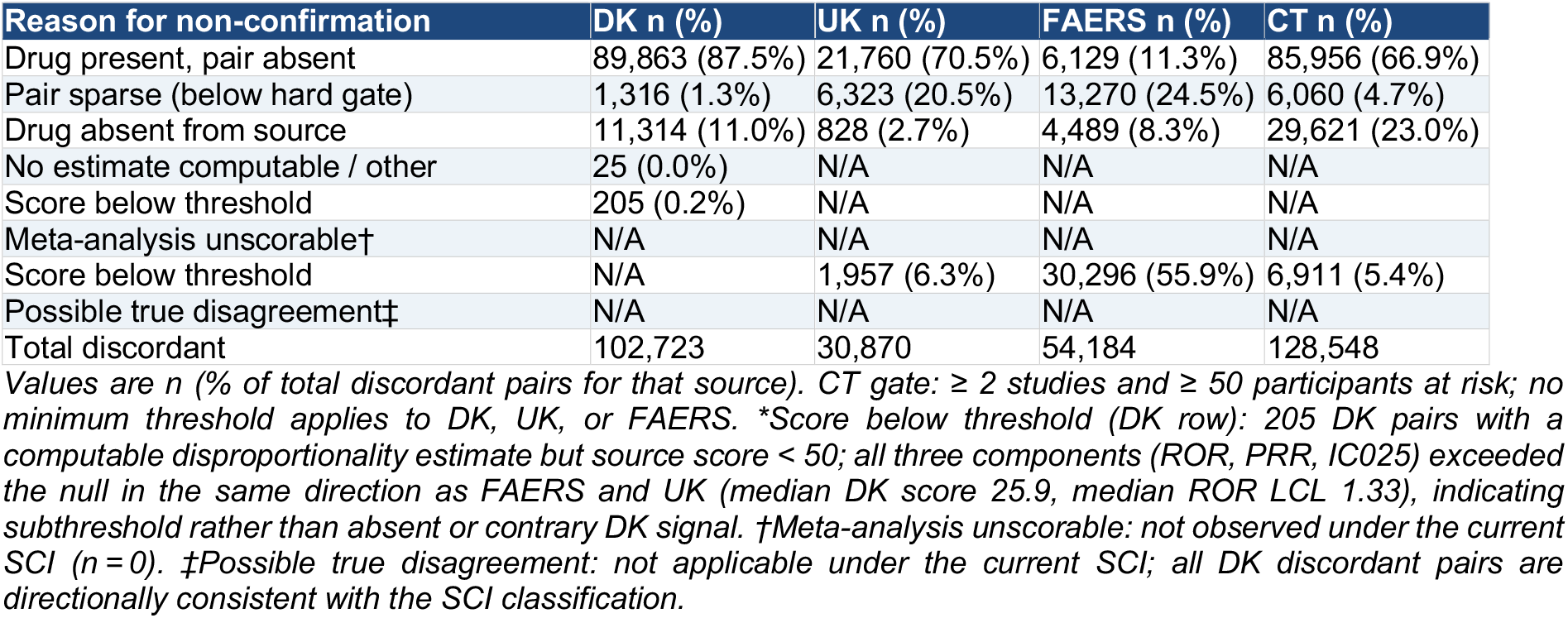
Source-specific discordance analysis: reasons for non-confirmation of SCI-positive signals by each individual source.

### 2.3 Temporal stability analysis

Across 5,861,548 drug-adverse event pairs with data in at least one temporal window, evaluated over eight cumulative data windows from 2015 to 2023, 98.1% received identical SCI classifications in every window in which they appeared (always-same), and 98.7% received the same classification in at least 80% of their windows (modal-dominant). This overall stability is partly structural: the large majority of pairs are non-signals in all windows. Among the 7,330 pairs classified as Strong in the final 2015-2023 window, the dominant pattern was progressive emergence rather than sudden appearance. Weak classifications were highly stable: 99.5% of 5,745,081 final-Weak pairs were always classified as Weak, and at most 0.5% reached Moderate or Strong in any individual window.

Strong signals demonstrated a characteristic progressive emergence pattern rather than sudden appearance (Table 4). Of the 7,330 pairs classified as Strong in the final 2015-2023 window, 6,573 (89.7%) had sufficient data for classification in the earliest 2015-2016 window; of these, 1,264 (17.2% of all 7,330 final-Strong pairs) were already classified as Strong, and 4,177 (57.0%) were already classified as Moderate or Strong. The remaining 757 pairs (10.3%) had no data in the first window, entering the detectable set as data accumulated in later years. Progressive emergence was the dominant pattern: by 2021, 5,592 of 7,330 final Strong signals (76.3%) were already classified as Strong, and 7,081 (96.6%) as Moderate or Strong. By 2022, 6,398 (87.3%) were Strong and 7,264 (99.1%) were Moderate or Strong. All 7,330 Strong signals reached Strong classification by the final 2015-2023 window. Classification stability once a Strong classification was first achieved was high: among the 500 most variable pairs in the dataset, only 2 of 433 final-Strong pairs showed a Weak classification after first achieving Strong (cefuroxime-cardiac arrest and donepezil-confusional state), both involving small evidence bases with transient source-level instability.

**Table 4.** Temporal stability of Strong signal classifications: classification of 7,330 final Strong signals across eight cumulative data windows from 2015 to 2023.

| Data window | Pairs with data | Classified Strong n (%) | Classified Moderate or Strong n (%) | Classified Weak n (%) | Not yet detectable n (%) |
| --- | --- | --- | --- | --- | --- |
| 2015-2016 | 6,573 | 1,264 (17.2%) | 4,177 (57.0%) | 2,396 (32.7%) | 757 (10.3%) |
| 2015-2017 | 6,904 | 1,965 (26.8%) | 5,017 (68.4%) | 1,887 (25.7%) | 426 (5.8%) |
| 2015-2018 | 7,096 | 2,663 (36.3%) | 5,722 (78.1%) | 1,374 (18.7%) | 234 (3.2%) |
| 2015-2019 | 7,185 | 3,413 (46.6%) | 6,195 (84.5%) | 990 (13.5%) | 145 (2.0%) |
| 2015-2020 | 7,269 | 4,088 (55.8%) | 6,546 (89.3%) | 723 (9.9%) | 61 (0.8%) |
| 2015-2021 | 7,317 | 5,592 (76.3%) | 7,081 (96.6%) | 236 (3.2%) | 13 (0.2%) |
| 2015-2022 | 7,329 | 6,398 (87.3%) | 7,264 (99.1%) | 65 (0.9%) | 1 (0.0%) |
| 2015-2023 | 7,330 | 7,330 (100.0%) | 7,330 (100.0%) | 0 (0.0%) | 0 (0.0%) |

*Pairs with data:* number of final-Strong pairs with at least one contributing source in that window. *Strong n (%)*: proportion of all 7,330 final-Strong pairs classified as Strong in that window. *M-or-S n (%)*: proportion classified as Moderate or Strong. *Weak n (%)*: pairs with data in that window but classified as Weak. *Not yet n (%)*: pairs with no data in that window across any source, computed as 7,330 minus Pairs with data.

These findings confirm that the Moderate classification is not noise but a genuine early detection state: 57.0% of final-Strong pairs were already detectable as Moderate or Strong in the first data window, and 96.6% had reached at least Moderate by 2021. The temporal stability of Weak classifications confirms that the SCI does not generate spurious transient signals: at most 0.5% of final-Weak pairs reached Moderate or Strong in any individual window. Among final-Moderate pairs (109,137), 47.8% received a modal-dominant classification (same class in ≥80% of their windows), consistent with borderline evidence levels that are appropriately sensitive to accumulating data.

### 2.4 Weight sensitivity analysis

A weight sensitivity analysis varying source weights across seven pre-specified scenarios demonstrated that SCI classifications are broadly robust to weight specification (Tables 5 and 6). Overall, 98.5% of all 6,905,874 drug-adverse event pairs received identical SCI classifications across all seven weight scenarios (perfectly stable), and 99.5% received the same classification in at least five of seven scenarios (mostly stable). The 103,833 pairs (1.5%) that received different classifications under different weight scenarios were concentrated in the Moderate baseline tier, consistent with their position near the signal threshold.

**Table 5.** Weight scenarios tested in the sensitivity analysis.

| Weight scenario | DK | UK | FAERS | CT | Rationale |
| --- | --- | --- | --- | --- | --- |
| Equal | 25% | 25% | 25% | 25% | No source preference; equal contribution |
| PV favoured | 20% | 30% | 30% | 20% | Spontaneous reporting emphasis |
| CT favoured | 20% | 20% | 20% | 40% | Clinical trial evidence emphasis |
| Europe favoured | 30% | 30% | 20% | 20% | European regulatory emphasis |
| FAERS favoured | 15% | 20% | 45% | 20% | US database dominance |
| DK favoured | 40% | 20% | 20% | 20% | Danish national registry emphasis |
| UK favoured | 20% | 40% | 20% | 20% | UK Yellow Card emphasis |

*Primary analysis uses equal weights (25% per source, i.e. simple mean of available source scores). All scenarios used the same source exclusion and weight normalisation logic as the primary analysis.*

**Table 6.** Weight sensitivity transition matrix: classification outcomes across all seven weight scenarios.

| Baseline classification | Classification under varied weights | N observations | Proportion |
| --- | --- | --- | --- |
| Strong | Strong | 47,961 | 94.0% |
| Strong | Moderate | 3,054 | 6.0% |
| Strong | Weak | 15 | 0.03% |
| Moderate | Moderate | 752,349 | 88.2% |
| Moderate | Weak | 78,115 | 9.2% |
| Moderate | Strong | 22,738 | 2.7% |
| Weak | Weak | 47,387,748 | 99.9% |
| Weak | Moderate | 49,138 | 0.1% |

*N observations: total classifications across all 6,905,874 pairs × 7 weight scenarios (N = 48,341,118 observations).*
*Proportion: fraction of classifications for each baseline tier receiving that outcome across weight scenarios.*
*Strong→Weak transitions occurred in 15 of 51,030 Strong-baseline classifications (0.03%) and are confined to scenarios with extreme de-emphasis of the dominant contributing source.*

Strong signal classifications were more robust than Moderate but less absolute than under the prior SCI specification: 64.6% of Strong-classified pairs under the primary equal-weight specification remained Strong under all seven alternative weight scenarios, and 94.9% remained Strong in at least five of seven scenarios. Of 51,030 Strong-baseline classifications across the seven scenarios, 94.0% remained Strong, 6.0% transitioned to Moderate, and only 0.03% (15 classifications) transitioned to Weak — confirming that Strong→Weak transitions are rare and confined to extreme weight configurations. The pairs that reclassified typically had signal evidence unevenly distributed across sources, such that de-emphasising a heavily contributing source reduced the composite below the Strong threshold. Weak signal classifications were highly stable (99.5% perfectly stable, 99.9% of classification observations remaining Weak), confirming that the absence of signal is not an artefact of weight choice. Moderate classifications showed the greatest sensitivity to weight variation (44.5% perfectly stable, 83.0% mostly stable), with 9.2% of classification observations transitioning to Weak and 2.7% to Strong, consistent with the inherent uncertainty of borderline signals. These results confirm that the Strong classification represents a robust finding that is largely independent of the specific weighting scheme applied, while acknowledging that the broader Strong tier under the updated SCI includes some signals sensitive to weight specification.

## Discussion

We have described the Signal Consensus Index, a composite pharmacovigilance signal detection score that integrates evidence from four independent data streams, two European national spontaneous reporting systems, the US FAERS database, and meta-analysed clinical trial results, into a single 0-100 score with associated confidence and classification tiers. Across 6,905,874 drug-adverse event pairs, the SCI generated 129,176 Moderate-or-Strong signals and 7,290 Strong signals, with reference set validation against the EU-ADR reference standard demonstrating 100.0% PPV and 100.0% specificity across all 88 classifiable pairs, and signal stability demonstrated across seven plausible weighting schemes. The principal finding of the validation analyses is that cross-source concordance substantially constrains unilateral signal classification, improving the prioritisation of signals with genuine multi-jurisdictional support over signals arising from database-specific artefacts. The SCI is designed as a prioritisation tool rather than a confirmatory test; the validation analyses demonstrate that its classifications are stable, internally consistent, and concordant with established pharmacological knowledge, rather than that they constitute confirmed causal associations.

The reference set validation confirms zero false positives across all 44 classifiable negative controls, with 100.0% specificity and PPV at both the Moderate-or-Strong and Strong thresholds. The sensitivity of 40.9% at the Moderate-or-Strong threshold reflects the conservative cross-source confirmation requirement rather than an inability to detect known associations: each false negative was attributable to identifiable data limitations including drug name fragmentation, withdrawn drugs, and insufficient case counts for rare outcomes. Five false negatives had SCI scores ≥ 50 but confidence scores of 25, indicating that a signal was detectable in a single source but lacked multi-source corroboration; the confidence gate correctly suppressed these to Weak. The zero false positive rate is a structural property of the cross-source confirmation requirement: no negative control pair achieved sufficient concordance across independent evidence streams to be classified as a signal, even where individual source scores were elevated. The practical implication is that Moderate-classified signals warrant pharmacological attention when data availability constraints explain the confidence penalty, as the signal score itself exceeds the detection threshold.

The single-source comparison provides the most direct evidence for the added value of cross-source aggregation. FAERS alone generated 1,438,246 signals at the ≥ 50 source-score threshold, approximately eleven-fold more than the 129,176 SCI Moderate-or-Strong signals. This finding replicates the precision-sensitivity trade-off documented by Candore et al. across single-source databases [4], but extends it to demonstrate that cross-source concordance operates as a substantial volume filter: 94.8% of FAERS disproportionality signals are not confirmed by the SCI, and the 74,992 that are confirmed represent a more tractable set for clinical prioritisation. The complementary finding, 54,184 SCI-positive signals absent from FAERS, of which 8.3% involve drugs not present in the US reporting system at all, demonstrates the added sensitivity of European source integration. These pairs represent pharmacovigilance signals for European-authorised medicines that would be systematically missed by any FAERS-only framework.

The discordance analysis reveals that the overwhelming majority of source-specific non-confirmation reflects data availability constraints or threshold effects rather than genuine pharmacological disagreement. For DK, UK, and clinical trial sources, the dominant reason for non-confirmation is simply that the drug-adverse event combination has not been reported or studied, not that the source contradicts the signal. For FAERS the pattern is balanced between data availability and subthreshold disproportionality scores. Among the 102,723 DK-discordant pairs, only 205 (0.2%) had a subthreshold DK score; all three disproportionality components (ROR, PRR, IC025) were positive for these pairs (median ROR lower CI bound 1.33), indicating a weak signal in the same direction as FAERS (median score 94) and UK (median score 91), consistent with insufficient Danish data volume rather than genuine disagreement. No source produces signals that contradict the SCI classification; discordance across all four sources is attributable to data availability differences or subthreshold evidence in the same direction. This pattern supports the validity of the SCI composite: when multiple sources agree, they are agreeing about a real pharmacological association; when they disagree, the disagreement is almost entirely explicable by data availability differences rather than conflicting evidence.

The weight sensitivity analysis demonstrates that SCI classifications are broadly robust to the precise specification of source weights, though the magnitude of robustness depends on the classification tier. Strong→Weak transitions are extremely rare (0.03% of Strong-baseline classifications across all seven scenarios, 15 of 51,030), and 95.0% of Strong signals retain their Strong classification in at least five of seven scenarios. Weak classifications are nearly perfectly stable (99.5% across all scenarios). The greater instability of Moderate classifications (44.6% perfectly stable, 83.0% mostly stable) is expected and appropriate: Moderate signals are by definition borderline, and sensitivity to weight specification at the margin is a feature, not a flaw, correctly identifying pairs where evidence is present but not robustly multi-source. The broader Strong tier under the updated SCI (7,290 pairs at baseline) includes some signals whose classification depends on specific source emphasis, which is a natural consequence of extending the Strong threshold beyond only the most heavily cross-source-corroborated signals.

Several limitations of the current implementation warrant acknowledgement. The SCI combines heterogeneous evidence types across two conceptually distinct domains. Source-specific scoring methods apply a common disproportionality estimand to all three spontaneous reporting sources (DK, UK, FAERS) and a meta-analytic risk ratio to clinical trials. These two estimand classes are not equivalent and their combination should be understood as a convergence of independent lines of pharmacological evidence for prioritisation purposes, not as a meta-analytic synthesis of a common underlying quantity. Exposure information is not accounted for in the spontaneous reporting components, and reporting biases, including notoriety bias, stimulated reporting, and indication confounding, cannot be fully eliminated by cross-source concordance alone. Drug name harmonisation across jurisdictions represents a core validity challenge for any cross-source pharmacovigilance platform. The valproate-hepatotoxicity pair in the original face validity set, and the amoxicillin clavulanate-acute liver injury pair in the EU-ADR reference set validation, illustrate the problem concretely: amoxicillin clavulanate appears as amoxycillin in UK/DK databases and amoxicillin in FAERS, splitting a genuine cross-source signal into two single-source signals each correctly held at Weak (confidence 25). Similarly, 7,348 UK Yellow Card reports for sodium valproate were not captured under the INN term valproic acid. In both cases the SCI behaves correctly under its own logic, but the root cause is upstream data harmonisation rather than the scoring algorithm. This is not an isolated case; FAERS data contain numerous brand name and salt form variants that may not map to INN terms used in European databases, and ClinicalTrials.gov intervention names are often formulation-specific. The cross-source concordance logic of the SCI depends on correct drug name matching across sources: a signal that appears as drug A in one source and drug B in another will not be recognised as concordant even if A and B are the same molecule. Systematic drug name normalisation, including INN mapping, salt form equivalence, and brand-to-generic linkage, is therefore a prerequisite for complete cross-source signal detection and a priority for future platform development. Signal detection is conducted at MedDRA Preferred Term level; higher-level groupings (High Level Terms, System Organ Classes) are not currently integrated, which may affect sensitivity for signals dispersed across related but non-identical preferred terms. The clinical trial meta-analysis component cannot produce a scorable estimate when all events occur in the treatment arm and control arms have zero events. Under the updated SCI this pattern is absorbed into the pair-sparse and subthreshold discordance categories rather than appearing as a separate “unscorable” class; the underlying issue, and the methodological opportunity it represents for alternative estimators of zero-event control arms, remain. The clinical trial gate thresholds (minimum two qualifying trials, minimum 50 participants at risk) represent pragmatic choices that could be varied; sensitivity analyses varying gate thresholds are planned for future work. The four data sources differ in data currency: UK Yellow Card and FAERS data extend through Q4 2025, while the Danish registry extends through 2023. This temporal imbalance means that signals emerging in 2024-2025 may be corroborated by UK and FAERS but cannot yet be confirmed by Danish data, potentially affecting confidence scores for recently emerging signals. As Danish data for 2024 onwards becomes available, classifications for borderline pairs may shift. This represents an inherent characteristic of real-time multi-source pharmacovigilance rather than a methodological flaw, and is shared by all platforms integrating data sources with different update cycles.

Future directions include extension of the temporal stability analysis to include annual granularity within each window, enabling more precise characterisation of signal emergence timing. Extension of the platform to incorporate EudraVigilance data would add a fifth independent European regulatory evidence stream. The SCI framework is designed to be drug-class and indication agnostic; a companion analysis applying the SCI to semaglutide demonstrates its utility for detecting both established adverse reactions and drug ineffectiveness signals in a high-volume contemporary therapeutic area (Jensen A, manuscript in preparation). As the platform matures, systematic comparison of SCI signal classifications against regulatory label updates would provide prospective evidence of clinical utility beyond the face validity demonstrated here.

## Data Availability

All data produced are available online at

https://aeatlas.com

